# Individual and School-Level Factors Associated with Pediatric Eye Disorders and Referral Adherence in an Enhanced School-Based Vision Screening Program in Ghana

**DOI:** 10.64898/2026.02.02.26345428

**Authors:** Afua O Asare, Priscilla Ablordeppey, Enoch A Asiedu, Emmanuel T Doku, Gabriel K Agbeshie, Seyram A Gle, Nana Akwasi O Mensah, Ruby E Adikah, Christine N Yeboah, Debora A Baidoo, Christine K Darko, Elisha E Arkhurst, Melissa H Watt, Eldrick A Acquah, Hornametor Afake, Sylvia Agyekum, Kwadwo O Akuffo

## Abstract

**Background:** Evidence on how social and school-level contexts shape pediatric vision screening outcomes is limited, particularly in sub-Saharan Africa. We examined the association between individual and contextual factors and vision screening outcomes in a pilot enhanced school-based vision screening program (ESVSP) in Kumasi, Ghana.

**Methods:** We conducted a cross-sectional study using data from an ESVSP to detect eye disorders in school-children aged 4 to 22 years. Outcomes were the presence of eye disorders and referral adherence. Exposure variables were individual [(age, sex, socioeconomic status (SES)], and contextual [school type (public vs private)] factors. Logistic regression was used to estimate unadjusted (OR) and adjusted (aOR) odds ratios with 95% confidence intervals (CI).

**Results:** We analyzed data for 1,123 children screened and 299 referred. The average age was 10.2 (±2.6) years. Overall, 34% (n=382) had suspected eye disorders, and 32.8% (n=98) adhered to the referral. After adjusting for key variables, children attending public (32.2%) compared to private (67.8%) schools had 45% lower odds of identified eye disorders (aOR= 0.55; 95% CI 0.37, 0.83). Children with low (13.3%) compared to high (28.6%) SES had 70% lower odds of referral adherence (aOR= 0.30; 95% CI 0.12, 0.80).

**Conclusion:** In this pilot school-based program, school context and socioeconomic status were associated with suspected pediatric vision and eye disorders, and referral adherence, respectively. These findings highlight equity-relevant gaps in referral adherence and underscore the need for context-specific strategies to strengthen referral pathways in low-resource settings.

## INTRODUCTION

The early years of childhood are crucial for the optimal development of the visual system, essential for early education, lifetime development, and quality of life.(1) Undetected and delayed management of pediatric eye disorders can result in permanent vision loss and disability. About 90% of childhood vision loss in Ghana is considered avoidable, with the leading cause of reduced vision being uncorrected refractive error (26% - 72%) and amblyopia (10%) from regional studies.(2-6)

Early detection of pediatric eye disorders is facilitated through vision screening.(7, 8) However, vision screening has limited value unless children identified with eye disorders complete referrals and receive needed treatment.(9-11) School-based vision screening programs in Ghana have reported the prevalence of eye disorders ranging between 1% and 8% based on the study population and screening methods used.(12-14)

Vision screening outcomes may be influenced by individual and contextual factors. For instance, individual factors such as socioeconomic status (SES), cultural factors, and social deprivation are associated with timely access to eye care and treatment adherence,(15-17) which subsequently impact diagnosis and severity of eye disorders.(18, 19) Contextual factors like environmental and school-related factors impact the development of eye disorders like refractive error among children. Time spent outdoors is protective against myopia,(20-24) and possibly astigmatism.(25, 26) However, evidence on how school context shapes these exposures and vision outcomes remains limited, particularly in Sub-Saharan Africa. Other contextual factors, such as the support of school administrators, may also contribute to the success of school-based vision screening programs.

Few studies have explored the influence of individual and particularly contextual factors on vision screening outcomes, especially among school children in low and middle-income countries.(2, 12, 15, 27-33) Targeted school-based vision screening could be a cost-effective way to bring vision care to the populations at highest risk of vision impairment. However, effective planning for these targeted programs in low and middle-income countries requires a thorough understanding of how individual and contextual factors influence vision screening outcomes, specifically the identification of eye disorders and adherence to eye care professional (ECP) referrals. This study aims to explore the association between individual and contextual factors and vision screening outcomes using data from a pilot enhanced school-based vision screening program (ESVSP) in Kumasi, Ghana.

## METHODS

### Study Design and Sample

This cross-sectional study evaluated a pilot ESVSP among children attending one public and one private school in Kumasi, Ghana to inform local implementation and planning. The two schools were located in the same neighborhood of Kumasi, 0.7 miles apart on foot, and 1.7 miles by car. The Ghana Education Service (GES) operates public schools in Ghana, while private schools are operated by private entities, providing curriculum developed by the GES, or other international curricula such as the British Cambridge education system.(34) In Ghana, public schools do not charge tuition fees for basic education under the government’s Free Compulsory Universal Basic Education (FCUBE) policy.(35) Nonetheless, families incur indirect costs such as uniforms, textbooks, and transportation, making public schools comparatively more accessible to children from low SES backgrounds than private schools. This study was designed to generate context-specific evidence to inform the implementation and scale-up of school-based vision screening programs in similar low-resource settings. The Committee on Human Research Publication, and Ethics at the Kwame Nkrumah University of Science and Technology, Kumasi, reviewed and accepted the study proposal (CHRPE/AP/866/24).

### Enhanced School-based Vision Screening Program

The ESVSP implemented from November to December 2024, was described as ‘enhanced’ due to the use of local optometrists and the inclusion of direct ophthalmoscopy. In many high-income countries, vision screening is carried out by trained laypersons or health care providers who are not ECPs (optometrist or ophthalmologist) and without direct ophthalmoscopy.(36-41) In our study, we were able to go beyond basic screening techniques by introducing direct ophthalmoscopy to identify abnormalities in the posterior segment of the eyes, such as glaucoma.

We defined an eye disorder as the presence of visually significant refractive error with an increased risk for amblyopia and ocular anomalies identified using the ESVSP protocol. Because complete eye health is not evaluated in vision screenings, definitive diagnosis is limited, unlike comprehensive eye exams.(42) For this reason, eye disorders detected during the vision screening were considered “suspected” and required a comprehensive examination for diagnosis.

All identified cases of suspected eye disorders were referred using a referral criterion developed by the American Academy of Pediatrics (AAP) and the American Academy of Pediatric Ophthalmology and Strabismus (AAPOS) to inform instrument-based screening (S1 Supplementary Table).(38, 43) Children were also referred if a two-line difference in visual acuity (VA) was observed between the two eyes, even when both eyes had VA within normal ranges. In addition, children were referred if they were unable to complete any of the screening tests after two attempts, including cases of malfunction of instrument-based cameras. Because of the high prevalence of glaucoma in Ghana,(44, 45) children with a suspicious disc defined as a Cup to Disk Ratio (CDR) ≥ 0.5, a CDR difference of 0.2 between each eye, or the presence of a laminar dot sign were also referred. S1 Supplementary Table describes the detailed referral criteria used in the ESVSP. For children with abnormal vision screening results, a referral note (with the vision screening findings) was sent home with the children to their guardians within 3 weeks of the screening. The referral note contained the address and phone numbers of two optometry clinics collaborating with the research team to facilitate appointment scheduling. These clinics were each within 3 miles (4.83 km) of each school. Guardians were informed that they could visit any eye clinic of their choice. They were also instructed to bring the referral note to the referral appointment, while optometrists were tasked to document the results of their comprehensive eye exams on a form attached to the referral note, which was later collected by the research team.

### Measures

The primary outcome variable for this study was the presence of an eye disorder (yes/no) after the vision screening test. Eye disorders were categorized as non-refractive (based on eye structure and function) and refractive eye disorders for descriptive purposes. The secondary outcome variable was referral adherence (yes/no) for children identified with an eye disorder after the vision screening test. The selection of exposure variables was informed by Andersen’s Behavioural Model of Health Services Use. Specifically, a contextual-level factor, which was the type of school (private or public) that the child was enrolled in (and where the vision screening test was done), and an individual-level factor, which was age, sex, and SES. All variables were assessed as categorical, except for age, which was assessed as a continuous variable.

Demographic variables, specifically, age, sex, and parents’ occupation, were collected from school enrollment records and in-person interviews with the children in June 2025. To enable the assessment of SES based on parents’ occupation, we adopted the International Standard Classification of Occupations (ISCO) framework to classify occupations into an organized hierarchy. The ISCO framework is a structured system developed by the International Labour Organisation (ILO) to classify jobs based on the tasks and duties performed. The framework facilitates cross-national comparisons of occupational data. The most recent version of the framework, ISCO-08 is widely used in social science research to derive occupational prestige scores and socioeconomic indices.(46) In this study, ISCO-08 codes were assigned to data collected on the occupations of both the father and mother of children who participated in the study using the University of Warwick’s Institute for Employment Research Computer Assisted Structured Coding Tool (CASCOT).(47) Data of children for whom both parents’ occupations were missing, as well as occupational information that could not be classified under any of the classifications in CASCOT were excluded from the sample data analyzed. Each ISCO-08 code was then converted using the *iscogen package* in StataNow/MP version 19.5 for Windows into a socioeconomic index; specifically, the *International Socio-Economic Index (ISEI) of Occupational Status*.(48) A higher ISEI index indicated higher social standing and access to resources based on one’s occupation (i.e., high SES) and vice versa.

### Statistical analysis

Stata (version 19.5, StataCorp, College Station, TX, US) was used to perform all analyses. Summary descriptive statistics, such as means and standard deviations, were computed for all continuous variables. Frequencies and percentages were computed for categorical variables. The Pearson chi-square test was used to assess the association between categorical exposures and the categorical outcome variables. To assess the association between the types of eye disorders and the study outcomes, the Fisher’s Exact test was used because of small cell sizes. Independent sample t-tests were used to assess the differences in continuous variables across categorical variables.

Multivariable logistic regression models were built to estimate the unadjusted odds ratio (OR), adjusted odds ratio (aOR), and 95% confidence interval (CI) for the association between the exposure and outcome variables. Each outcome modeled was adjusted for sex, age (years), type of school, and SES. Statistical significance for all analyses was defined by *p* < .05

## RESULTS

### Eye Disorders and Factors Associated

Data was analyzed for the 1,123 children screened (Table I). There were 686 (61.1%) children who attended private school, 341 (30.4%) with low SES, and 648 (57.7%) were female. The average age of children screened was 10.2 (±2.6) years. Of the children screened, 382 (34.0%) were detected to have a possible eye disorder. Eye disorders were observed more frequently among children attending the private school 259 (67.8%), children with moderate SES [118 (30.9%], as well as females [202 (52.9%)]. The average age of children detected with an eye disorder was 10.3 (±2.7) years. 156 (13.9%) screened children had refractive errors (S2 Supplementary Table), which were observed more frequently in private school [120 (76.9%)]. Non-refractive errors were also commonly identified in private school [158 (64.8%)] (S3 Supplementary Table). There were 26 (2.3%) screened children with myopia (S4 Supplementary Table), with the condition being predominant in private schools [24 (92.3%)]. Astigmatism was identified in 82 (7.3%) children (S5 Supplementary Table) and was more frequently observed in private schools [65 (79.3%)]. Children attending the public school had lower odds of having an eye disorder compared with children from the private school before (OR 0.65; 95% CI 0.50, 0.84) and after (aOR 0.55; 95% CI 0.37, 0.83) adjusting for SES, sex, and age (Table 2). Descriptions of all eye disorders detected by type of school and referral adherence are in Supplementary materials S6-S8.

**Table 1.** Descriptive characteristics of children completing school-based vision screening and the presence of a suspected eye disorder.

| Characteristic | Total<br>(n=1,123) | Presence of Eye Disorder (n, %) |  | p-value |
| --- | --- | --- | --- | --- |
|  |  | Yes<br>(n=382) | No<br>(n=741) |  |
| School Type |  |  |  |  |
| Public | 437 (38.91) | 123 (32.20) | 314 (42.38) | 0.001 |
| Private | 686 (61.09) | 259 (67.80) | 427 (57.62) |  |
| Socioeconomic Status |  |  |  |  |
| Low | 341 (30.37) | 107 (28.01) | 234 (31.58) | 0.101 |
| Moderate | 303 (26.98) | 118 (30.89) | 185 (24.97) |  |
| High | 264 (23.51) | 93 (24.35) | 171 (23.08) |  |
| Missing | 215 (19.15) | 64 (16.75) | 151 (20.38) |  |
| Sex |  |  |  |  |
| Male | 475 (42.30) | 180 (47.12) | 295 (39.81) | 0.019 |
| Female | 648 (57.70) | 202 (52.88) | 446 (60.19) |  |
| <sup>a</sup> Age, years | 10.24 (2.60) | 10.26 (2.66) | 10.24 (2.58) | 0.879 |

<sup>a</sup> Age is represented by the mean ( ± standard deviation) for n=1,115. 8 children were excluded because of missing data. p-value based on an independent sample t-test

**Table 2.** Association between exposure variables and the presence of eye disorders identified in the vision screening study.

| Exposure | Presence of an Eye Disorder |  |  |  |
| --- | --- | --- | --- | --- |
|  | OR (95%) | p-value | aOR (95%) | p-value |
| School Type |  |  |  |  |
| Public | 0.80 (0.71, 0.91) | 0.001 | 0.55 (0.37, 0.83) | 0.004 |
| Private | Reference | - | Reference | - |
| Socioeconomic Status |  |  |  |  |
| Low | 0.84 (0.60, 1.18) | 0.318 | 1.29 (0.82, 2.05) | 0.272 |
| Moderate | 1.17 (0.83, 1.65) | 0.361 | 1.28 (0.90, 1.81) | 0.171 |
| High | Reference | - | Reference | - |
| Sex |  |  |  |  |
| Male | 1.35 (1.05, 1.73) | 0.019 | 1.31 (0.99, 1.73) | 0.058 |
| Female | Reference | - | Reference | - |
| Age, years | 1.00 (0.96, 1.05) | 0.878 | 1.04 (0.99, 1.10) | 0.123 |

OR = odds ratio; aOR = adjusted odds ratio

### Referral Adherence and Factors Associated

Data on referral adherence were available for 299 (78.3%) of the 382 children referred; 98 children adhered to the referral, indicating an overall referral adherence rate of 32.8% (Table 3). The rates of referral adherence for public and private schools were 18.8% and 39.4%, respectively. Of the children referred, 203 (67.9%) attended private school, 89 (29.8%) had low SES, 157 (52.5%) were females, and the average age for referral was 9.8 (±2.5) years. Referral adherence was more frequently observed among children in private school [80 (81.6%)], children with moderate SES [35 (35.7%]), as well as females [51 (52.0%)]. The average age of children who adhered to a referral was 9.4 (±2.5) years. Children with low SES had a lower odds of referral adherence compared to those with high SES before (OR 0.26; 95% CI 0.12, 0.56) and after (aOR 0.30; 95% CI 0.12, 0.80) adjusting for school type, sex, and age (Table 4).

**Table 3.** Descriptive characteristics of children referred for comprehensive eye examination and adherence rates.

| Characteristic | Total<br>(n=299) | Referral Adherence (n, %) |  | p-value |
| --- | --- | --- | --- | --- |
|  |  | Yes<br>(n=98) | No<br>(n=201) |  |
| School Type |  |  |  | <0.001 |
| Public | 96 (32.11) | 18 (18.37) | 78 (38.81) |  |
| Private | 203 (67.89) | 80 (81.63) | 123 (61.19) |  |
| Socioeconomic Status |  |  |  | <0.001 |
| Low | 89 (29.77) | 13 (13.27) | 76 (37.81) |  |
| Moderate | 93 (31.10) | 35 (35.71) | 58 (28.86) |  |
| High | 71 (23.75) | 28 (28.57) | 43 (21.39) |  |
| Missing | 46 (15.38) | 22 (22.45) | 24 (11.94) |  |
| Sex |  |  |  | 0.104 |
| Female | 157 (52.51) | 51 (52.04) | 106 (52.74) |  |
| Male | 142 (47.49) | 47 (47.96) | 95 (47.26) |  |
| <sup>a</sup> Age, years | 9.80 (2.53) | 9.36 (2.48) | 9.98 (2.53) | 0.048 |

<sup>a</sup> Age is represented by the mean ( ± standard deviation) for n=297 because of missing data. p-value based on an independent sample t-test

**Table 4.** Association between exposure variables and referral adherence in the vision screening study.

| Exposure | Referral Adherence |  |  |  |
| --- | --- | --- | --- | --- |
|  | OR (95%) | p-value | aOR (95%) | p-value |
| School Type |  |  |  |  |
| Public | 0.60 (0.44, 0.80) | 0.001 | 0.88 (0.56, 1.36) | 0.555 |
| Private | Reference | - | Reference | - |
| Socioeconomic Status |  |  |  |  |
| Low | 0.26 (0.12, 0.56) | 0.001 | 0.30 (0.12, 0.80) | 0.016 |
| Moderate | 0.93 (0.49, 1.75) | 0.814 | 0.98 (0.51, 1.86) | 0.948 |
| High | Reference | - | Reference | - |
| Sex |  |  |  |  |
| Male | 1.03 (0.63, 1.67) | 0.910 | 0.97 (0.55, 1.71) | 0.927 |
| Female | Reference | - | Reference | - |
| Age, years | 0.90 (0.82, 1.00) | 0.049 | 0.90 (0.79, 1.01) | 0.082 |

OR = odds ratio; aOR = adjusted odds ratio

## DISCUSSION

### Main findings of this study

This study explored the influence of individual and contextual factors on the presence of eye disorders and referral adherence using an ESVSP in Kumasi, Ghana. Children enrolled in the public school had lower odds of being identified with an eye disorder, compared to children enrolled in the private school, after adjusting for SES, age, and sex. Also, children with low compared to high SES had lower odds of referral adherence. The study results reveal significant disparities in the presence of eye disorders between school types and referral adherence based on SES for children participating in an ESVSP. School context and socioeconomic status may be important considerations in designing equitable, sustainable school-based vision screening and referral strategies in similar settings.

### What is already known on this topic

Prior studies exploring associations between school type and eye disorders in Nigerian and Ghanaian schools had inconsistent results, which may result from variations in school curricula, vision screening protocols, diagnostic criteria, study population, and confounding factors.(12, 27, 28, 30) We found that children attending the public school had a significantly lower odds of having an eye disorder compared to children in the private school after adjusting for SES, sex, and age. Though data is limited on African children, studies mostly in Asian populations have demonstrated that increased time outdoors is protective against the onset of myopia(21, 49-53) and astigmatism.(25, 26, 54) The higher prevalence of refractive errors, particularly myopia, among children attending the private school in our study may be consistent with differences in time spent outdoors and engagement in near work described in prior studies.(21, 49-53) Prior studies have reported higher levels of after-school tutoring in private schools, which has been hypothesized to increase near-work exposure.(55-57) Under the FCUBE policy, public schools are more accessible to children from lower socioeconomic backgrounds. Prior studies suggest that differences in daily activities among children, including outdoor exposure and near-work demands, may vary by socioeconomic status.(58) In the Ghanaian context, daily activities may differ by socioeconomic position and school context; however, these exposures were not directly measured in this study. These contextual differences underscore the importance of designing public health interventions that are sensitive to differences across school settings.

In this study, children with low compared to high SES had significantly lower odds of adhering to ECP referrals after adjusting for school type, age, and sex. Prior studies conducted in Israel,(59) India,(60) and Germany (61) have reported inconsistent findings regarding the relationship between individual and contextual factors and adherence to ECP referrals following abnormal vision screening tests. Such variability may stem from differences in how SES was measured across studies. To the best of our knowledge, no research in Sub-Saharan Africa has quantitatively examined the influence of SES on referral adherence after abnormal vision screening tests. However, earlier studies of Ghanaian school children have reported no significant association between SES and history of eye examinations.(12, 31) Addressing these barriers is essential for ensuring that the benefits of early detection translate into effective treatment and reduced inequities in child eye health.

Children with low SES face multiple barriers to accessing comprehensive vision care, including limited financial resources, parental misconceptions about the need for or effectiveness of treatment, and logistical challenges such as lack of transportation.(19, 62-66) In Ghana, low SES children living in informal foster arrangements or working as domestic workers often experience unstable caregiving and limited parental advocacy, which further reduces their access to preventive care and adherence to health referrals.(67) These children also encounter heightened economic and social vulnerability, as both they and their employers, who frequently serve as de facto guardians, may deprioritize health needs in favor of work responsibilities, resulting in missed appointments and untreated conditions. (68)

### What this study adds

This study provides context-specific evidence from a low-resource setting showing that both school context and socioeconomic status are associated with pediatric vision screening outcomes and referral adherence. By using standardized referral criteria and a validated internationally-recognized socioeconomic classification framework, the findings support cross-context comparisons and highlight actionable gaps in follow-up care that are relevant to global efforts to reduce avoidable childhood vision impairment.

### Limitations of this study

Despite its strengths, the study has some limitations. We performed non-cycloplegic refraction using the Plusoptix photoscreener, which may have overestimated myopia and underestimated hyperopia.(69-71) This could exaggerate school-type differences in myopia and attenuate differences in hyperopia without reversing the direction of associations. Also, the higher prevalence of myopia in our study should be interpreted cautiously because of the small number of cases. While the use of the ISCO framework for SES ranking is an overall strength of the study, there are some related limitations. Some occupations could not be classified using CASCOT. Another limitation is the inclusion of only two schools and the use of purposive instead of random sampling in the selection of schools and types. This approach may have introduced selection bias and a lack of generalizability to other populations. However, as a pilot study, findings are intended to inform local program planning rather than population-level estimates. As with many pilot evaluations, findings should be interpreted in light of limited generalizability but remain valuable for informing implementation in similar contexts. Although SES data were missing for 19% of children, missingness was observed more frequently in the public school, but was not associated with the study outcomes and did not bias our findings. Lastly, because referral adherence was defined by interviews with children and returned referral and parent survey forms, true adherence is likely underestimated among children who adhered to care but did not return forms.

## CONCLUSION

School-based vision screening programs are widely used to support early identification of eye disorders. However, their population-level impact is often limited by low referral completion following abnormal screening results. Previous research indicates that socioeconomic and contextual factors influence access to eye care, though evidence from Sub-Saharan Africa remains limited.

This study provides comparative evidence from two schools in the same urban neighborhood in Ghana, demonstrating that both school context and socioeconomic status are associated with vision screening outcomes. Using an enhanced screening approach with standardized referral criteria, children attending the public school had lower odds of being identified with suspected eye disorders, while children from lower socioeconomic backgrounds had lower odds of adhering to referrals. These findings illustrate how school context and socioeconomic disadvantage may contribute to inequities in pediatric eye health and impede the effectiveness of vision screening programs in low-resource settings.

## Declaration of Generative AI and AI-assisted technologies in the writing process

Generative AI and AI-assisted technologies were used in a limited and supportive manner during the preparation of this application to assist with editing for clarity, organization, and readability. AI technologies were not used to generate scientific ideas, formulate research questions, design study aims, interpret data, or make analytic decisions. The candidate reviewed, edited, and verified all content to ensure accuracy, originality, and alignment with the proposed research. Responsibility for the final content rests entirely with the candidate.

## Data availability

The data set associated with this paper is available upon request from the corresponding author.

## Ethics approval and Consent to participate

The Committee on Human Research, Publications and Ethics of the Kwame Nkrumah University of Science and Technology, Kumasi (KNUST), reviewed and approved the study protocol (Approval No. CHRPE/AP/866/24). The study was conducted in accordance with the principles of the Declaration of Helsinki. Written informed consent to participate was obtained from the parents or legal guardians of all participating schoolchildren.

## Author Contributions statement

Authors’ contributions: AOA, KOA, PA, EAA conceptualized and designed the study, acquired, analyzed data, and drafted the original article. ETD, GKA, SAG, NAOM, CNY, REA, DAB, CKD, EEA, Eldrick AA, SAA acquired the data, made substantial contributions to conceptualizing and designing the study, and revised it critically for important intellectual content. MHW, SV, AMH made substantial contributions to conceptualizing and designing the study, revising the manuscript critically for important intellectual content. All authors contributed to interpreting the data, approved the manuscript version to be published and agreed to be accountable for all aspects of the work.

